# Prevalence and Correlates of Symptoms of Cannabinoid Hyperemesis Syndrome in the United States

**DOI:** 10.64898/2026.01.25.26344780

**Authors:** Mark A. Ilgen, Amanda M. Price, Paula Goldman, Blair J. Whittington, Brian M. Hicks

**Affiliations:** Addiction Center, Department of Psychiatry, University of Michigan, Ann Arbor, MI; Center for Clinical Management Research, Department of Veterans Affairs, Ann Arbor VA Healthcare System, Ann Arbor, MI; Department of Pediatrics, University of Michigan, Ann Arbor, MI

## Abstract

**Objectives:** To determine the national prevalence of Cannabinoid Hyperemesis Syndrome (CHS) symptoms and associated characteristics.

**Methods:** Using data from a nationally representative survey of 7,034 US adults (conducted May-September 2025), we fit separate survey-weighted multinomial logistic regression models with a four-category CHS symptom and cannabis use group variable as the outcome and demographic characteristics, cannabis use behaviors, and cannabis-related problems as predictors.

**Results:** Overall, 2.7% of all respondents and 17.8% of respondents who used cannabis daily reported CHS-like symptoms. Respondents who were younger, female, identified as non-White, had lower income, had lower educational attainment, and endorsed cannabis-related use problems were more likely to be in the CHS symptom group than in the daily cannabis use group.

**Conclusions:** CHS symptoms are not uncommon in the US and those with fewer economic resources and more cannabis-related use problems were more likely to report these symptoms, even compared to others with daily cannabis use.

**Policy implications:** As cannabis use increases, CHS is also likely to become more common, underscoring the importance of expanded education about the identification and treatment of CHS.

## Introduction

Increasing legalization of cannabis in the United States (US) over the last 10 years has coincided with substantial increases in the prevalence of cannabis use.^1, 2^ There has been a more pronounced increase in heavy cannabis use, typically defined as daily or near-daily frequency, among US adults, which is estimated to have increased 269% between 2008 and 2022.^3^ As a potential consequence of heavy cannabis use, Cannabinoid Hyperemesis Syndrome (CHS) was first reported in 2004,^4^ and is characterized by recurrent episodes of severe nausea, vomiting, and abdominal pain among people who use cannabis frequently and for an extended period.^5–10^ The causes of CHS are not fully understood but are hypothesized to reflect the biphasic effects of cannabinoids, where infrequent administration of cannabis may produce antiemetic effects, while frequent, high dose exposure may overstimulate and dysregulate the vomiting control system in the brain.^6, 11–17^ In terms of strategies to improve CHS symptoms, many individuals report reduced nausea after hot showers or baths.^18^ The only effective long-term strategy for preventing future episodes appears to be cessation of cannabis use.^9, 13, 19^

Most of the existing literature on CHS is based on reviews of medical records from settings that treat patients during acute episodes of CHS. Recent data from emergency departments indicates that CHS rates have been rising alongside increased cannabis use, particularly in states where cannabis is legal.^20, 21^ Estimates from a large-scale retrospective study suggest that CHS may affect up to 6% of people evaluated for recurring vomiting within the Emergency Department,^22^ with hospitalizations and outpatient visits for related symptoms becoming more common in recent years.^23–25^ However, the existing literature is mostly limited to individuals who seek care for CHS episodes in medical settings, which does not include or reflect the broader population of individuals with heavy cannabis use.

National survey data are necessary to understand the overall frequency outside of medical settings and to understand the potential public health impact of CHS symptoms among those who use cannabis. We utilized data from a large nationally representative survey in 2025 of US adults to document potential symptoms of CHS and how these relate to differing patterns of cannabis use and cannabis-related problems, and general correlates of CHS symptoms.

## Methods

### Study Design

Data were collected as part of the National Firearms, Alcohol, Cannabis, and Suicide (NFACS) survey (N=7,034), a cross-sectional, nationally representative survey of US adults aged 18 years or older. The survey was developed by the study team and fielded by a market research company, Verasight, from May 27 – September 2, 2025. Participants were recruited via random address-based sampling (ABS) or multimedia messaging service (MMS) and single messaging service (SMS) text message methods; recruitment is depicted in Figure 1.

For the ABS sample, potential participants were identified through the US Postal Service database, which includes all non-institutionalized US adults with a current mailing address. Individuals selected from the sample were mailed an initial invitation letter as well as a reminder postcard. Where corresponding cell phone numbers and email addresses were available, the research company also sent follow-up reminders with a direct URL to the web-based survey utilizing those alternative contact methods. A total of 49,988 initial invitations were sent for ABS recruitment, with 1,914 responses received (3.83% response rate). From these responses, a total of 306 (15.99%) were removed for being incomplete, and an additional 99 responses (6.16%) were excluded during the quality assurance process, which included confirming all responses corresponded with a US IP address, no duplicate responses were provided, and no non-human responses were included. Participants who completed the survey in less than 30% of the median completion time were also removed. This produced a total of 1,509 participants.

For the MMS and/or SMS text message recruitment, potential participants were identified from a database consisting of voter file and supplemental commercial records and received a survey link via text message. A total of 2,227,925 invites were sent via MMS and SMS text messages, with 8,652 responses received (0.39% response rate). Twenty-nine percent (n=2,523) of those responses were removed for incomplete data, and an additional 9.85% (n=604) were removed for quality control, resulting in an MMS/SMS sample of 5,525 participants. Responses were weighted to match national benchmarks from the July 2025 Current Population Survey for age, sex, race and ethnicity, income, education, political party identification, region, and metropolitan status.

The protocol was reviewed by the University of Michigan Health Sciences and Behavioral Sciences IRB and received an exempt determination. Prior to accessing the survey, participants reviewed information on the survey content, length of the survey and compensation, and were informed that participation was voluntary and anonymous. All participants provided active consent prior to receiving study questions and all participants completed the survey online.

### CHS symptoms

Several sequential questions were used to capture a broad group of respondents who endorsed symptoms consistent with CHS. First, all respondents who endorsed at least 20 lifetime uses of cannabis were asked if they had a period of daily or near daily cannabis use in the past 5 years, which coincides with increased cannabis legalization and availability of high potency cannabis products. Those who endorsed daily use in the past 5 years were asked if they had experienced periods of severe nausea, vomiting, or abdominal pain. Those who endorsed such periods were asked about frequency and if they received symptom relief from taking hot baths or showers or from discontinuing cannabis use for a long period of time. Finally, all respondents who endorsed at least 20 lifetime uses of cannabis were asked whether they have ever received a diagnosis of CHS. It is important to reiterate that these items were generated by our research group as a novel way to measure potential CHS symptoms and have not been validated in other studies.

Based on this information, we made four groups for comparisons: the <u>*CHS symptom group*</u> consisted of respondents who endorsed a period of daily cannabis use in the past 5 years and periods of nausea, vomiting, or abdominal pain (*n* = 191); the <u>*daily cannabis use group*</u>, consisted of all other respondents who endorsed a period of daily cannabis use in the past 5 years but not periods of severe nausea, vomiting, or abdominal pain (*n* = 882); the <u>*cannabis use in the past 12 months group*</u> consisted of all other respondents who reported cannabis use in the past 12 months (*n* = 1288); the <u>*no cannabis use in the past 12 months group*</u> included all other respondents (*n* = 4673).

### Cannabis use and problem use

Lifetime and past 12 month cannabis use was assessed using the Alcohol, Smoking, and Substance Involvement Screening Test (ASSIST).^26^ Problem use was measured through the presence of negative consequences of cannabis use from items similar to the Cannabis Use Disorder Identification Test-Revised (CUDIT-R),^27^ with some additions including items that assessed lifetime problem use as well as past 12-month prevalence. In addition to specific cannabis use problems, we calculated a continuous latent trait score of cannabis problem use by fitting a 2-parameter item response theory model to the cannabis problem use items (separately for the lifetime and past 12 months items).

### Analyses

We used poststratification survey weights to estimate the prevalence of cannabis use and CHS symptoms. To examine demographic differences across the CHS symptom and cannabis use groups, we fit separate unadjusted multinomial logistic regression models, with the four-category CHS symptom and cannabis use group variable as the outcome and each demographic characteristic as the predictor. To compare groups on past 12-month cannabis use behaviors and lifetime cannabis use problems, we fit separate unadjusted and adjusted multinomial logistic regression models. In these models, the four-category CHS symptom and cannabis use group variable was the outcome, and each cannabis use behavior or cannabis use problem was the predictor. Adjusted multivariable models included age, sex, race and ethnicity, income, and education as covariates. In post hoc analyses, we fit separate unadjusted and adjusted logistic regression models using binary group indicators for pairwise comparisons of the CHS symptom group with each of the other cannabis use groups as the outcomes to assess differences on the predictors of interest. All analyses were conducted in Stata 19 using poststratification weights to account for complex survey design.

## Results

### Prevalence of CHS symptoms

Table 1 presents prevalence estimates for cannabis use and CHS symptoms. Approximately one-third of respondents reported cannabis use in the past 12 months, and 15.3% reported a period of daily use in the past 5 years. Overall, 2.7% of respondents reported episodes of severe nausea, vomiting, or abdominal pain, and 0.44% reported receiving a medical diagnosis of CHS. Among adults who had ever used cannabis, 5.2% reported CHS symptoms. Among respondents with a period of daily cannabis use in the past 5 years, 17.8% reported CHS symptoms, of whom 11.5% reported receiving a medical diagnosis of CHS. Among respondents who reported CHS symptoms, approximately two-thirds indicated that episodes of nausea, vomiting, or abdominal pain occurred monthly or less often, whereas the remaining one-third indicated these symptoms occurred weekly or more frequently. Nearly half of those endorsing CHS symptoms reported symptom relief by taking hot baths or showers. Only one-quarter indicated that stopping cannabis use improved symptoms, though the items did not assess how many respondents with CHS symptoms attempted to abstain from cannabis use.

**Table 1.** Prevalence of cannabis use and symptoms of cannabinoid hyperemesis syndrome; National Firearms, Alcohol, Cannabis, and Suicide survey, 2025.

|  | *n* | Percent [95% CI] | Population estimate [95% CI] |
| --- | --- | --- | --- |
| Cannabis use |  |  |  |
| Past 12 months | 2305 | 32.77 [31.29, 34.28] | 86,670,172<br>[82,265,905 – 91,074,438] |
| Period of daily or almost daily cannabis use in<br>the past 5 years | 1072 | 15.25 [14.09, 16.48] | 40,327,049<br>[37,263,940 – 43,593,656] |
| Period of daily use and episodes of severe nausea,<br>vomiting, or abdominal pain | 191 | 2.71 [2.20, 3.34] | 7,179,961<br>[5,662,092 – 8,697,830] |
| Diagnosed with cannabinoid hyperemesis<br>syndrome | 31 | 0.44 [0.23,0.83] | 1,155,788<br>[416,444 – 1,895,133] |
| Percent daily users with episodes of severe<br>nausea, vomiting, or abdominal pain | 191 | 17.80 [14.65, 21.47] |  |
| Frequency of symptoms |  |  |  |
| 1 x year | 50 | 26.28 [18.67, 35.65] |  |
| 1 x month | 79 | 41.45 [31.04, 52.69] |  |
| 1 x week | 24 | 12.33 [7.16, 20.40] |  |
| More than weekly | 38 | 19.93 [12.88, 29.53] |  |
| Symptoms improved..... |  |  |  |
| After stopping cannabis use | 53 | 27.59 [18.48, 39.04] |  |
| By taking hot baths or showers | 90 | 47.16 [36.63, 57.94] |  |
| Either stopping cannabis use or taking hot baths<br>or showers | 105 | 54.83 [44.23, 65.02] |  |

Note. Number of observations, proportions, and population estimates were estimated using poststratification weights to match national benchmarks of the July 2025 Current Population Survey for age, sex, race and ethnicity, income, education, political party affiliation, region, and metropolitan status for adults (i.e., 18 years or older) living in the United States (264,498,611).

### Demographic differences

Table 2 presents prevalence estimates and unadjusted multinomial regression results examining demographic differences across the four CHS symptom and cannabis use groups. Relative to the daily cannabis use group, younger age was significantly associated with being in the CHS symptom group, whereas older age was significantly associated with being in the no cannabis use in the past-12 months group. Non-White respondents were also more likely than White respondents to be in the CHS symptom group rather than the daily cannabis use group. In contrast, race and ethnicity was not significantly associated with being in either the cannabis use in the past 12-months or the no cannabis use in the past 12 months groups versus the daily cannabis use group. Relative to the daily cannabis use group, lower income and lower educational attainment were significantly associated with greater likelihood of being in the CHS symptom group, but lower likelihood of being in the past 12-month cannabis use and no past 12-month cannabis groups. Post hoc logistic regression models indicated that the CHS symptom group differed significantly from both the past 12-month cannabis use and no cannabis use in the past 12-months groups on all demographic characteristics, except sex.

**Table 2.**
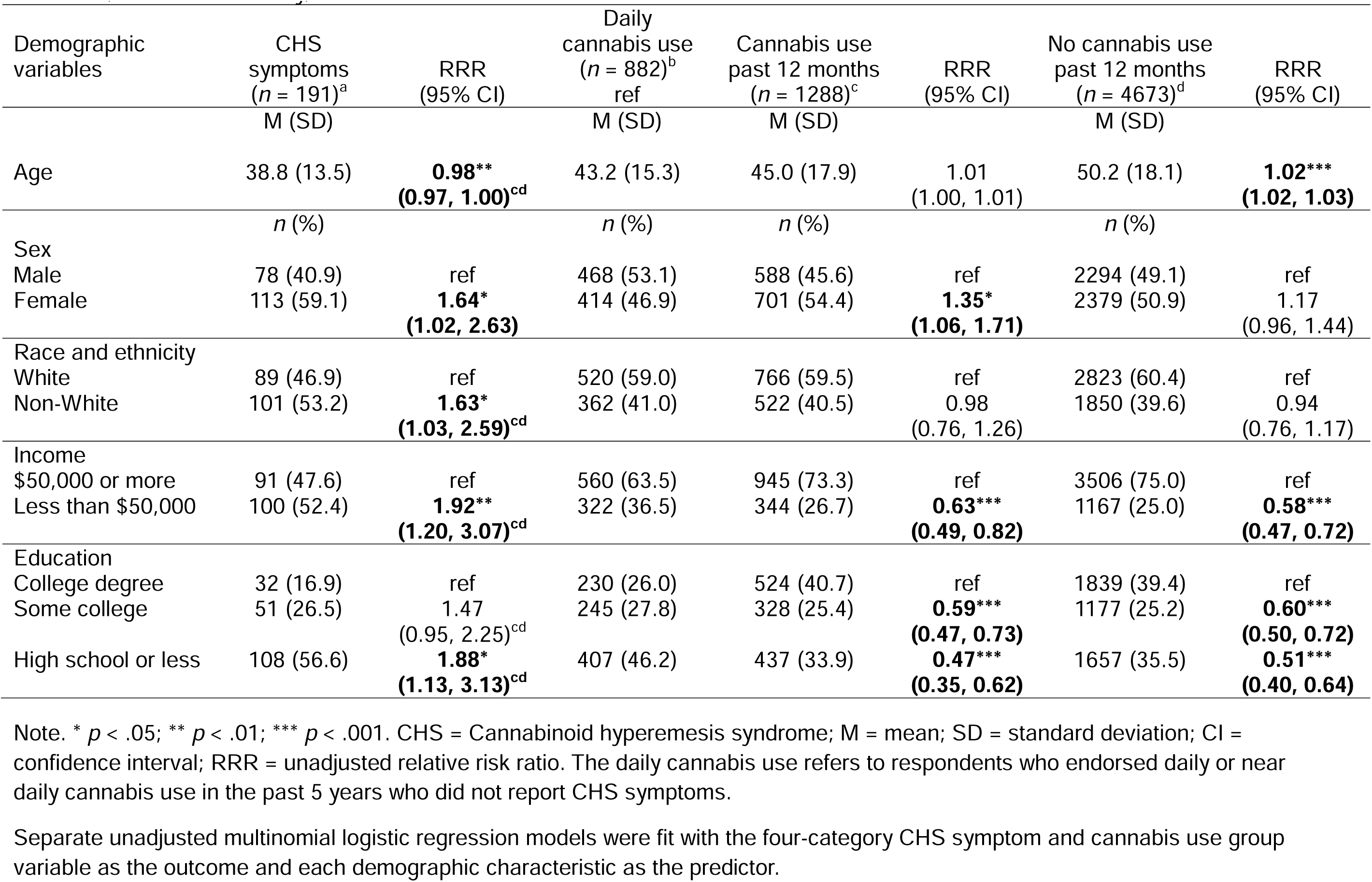
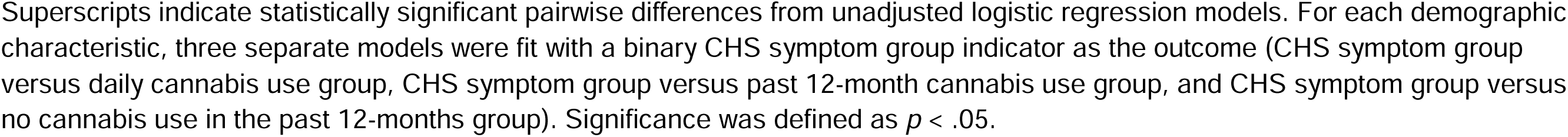
Demographic differences among cannabinoid hyperemesis syndrome and cannabis use groups; National Firearms, Alcohol, Cannabis, and Suicide survey, 2025.

### Cannabis use

As seen in Table 3, the prevalence of being high or stoned 7 or more hours on a typical day when using cannabis was 24.4% in the CHS symptom group and 15.1% in the daily use group, compared with 3.2% in the cannabis use in the past 12 months group. There was not a significant difference in the rate of being high or stoned for 7 or more hours on a typical day between the CHS symptom group and the daily cannabis use group in either unadjusted or adjusted models. However, the CHS symptom group and daily cannabis use group reported significantly higher rates being high or stoned for 7 or more hours than the cannabis use in the past 12-months group.

**Table 3.**
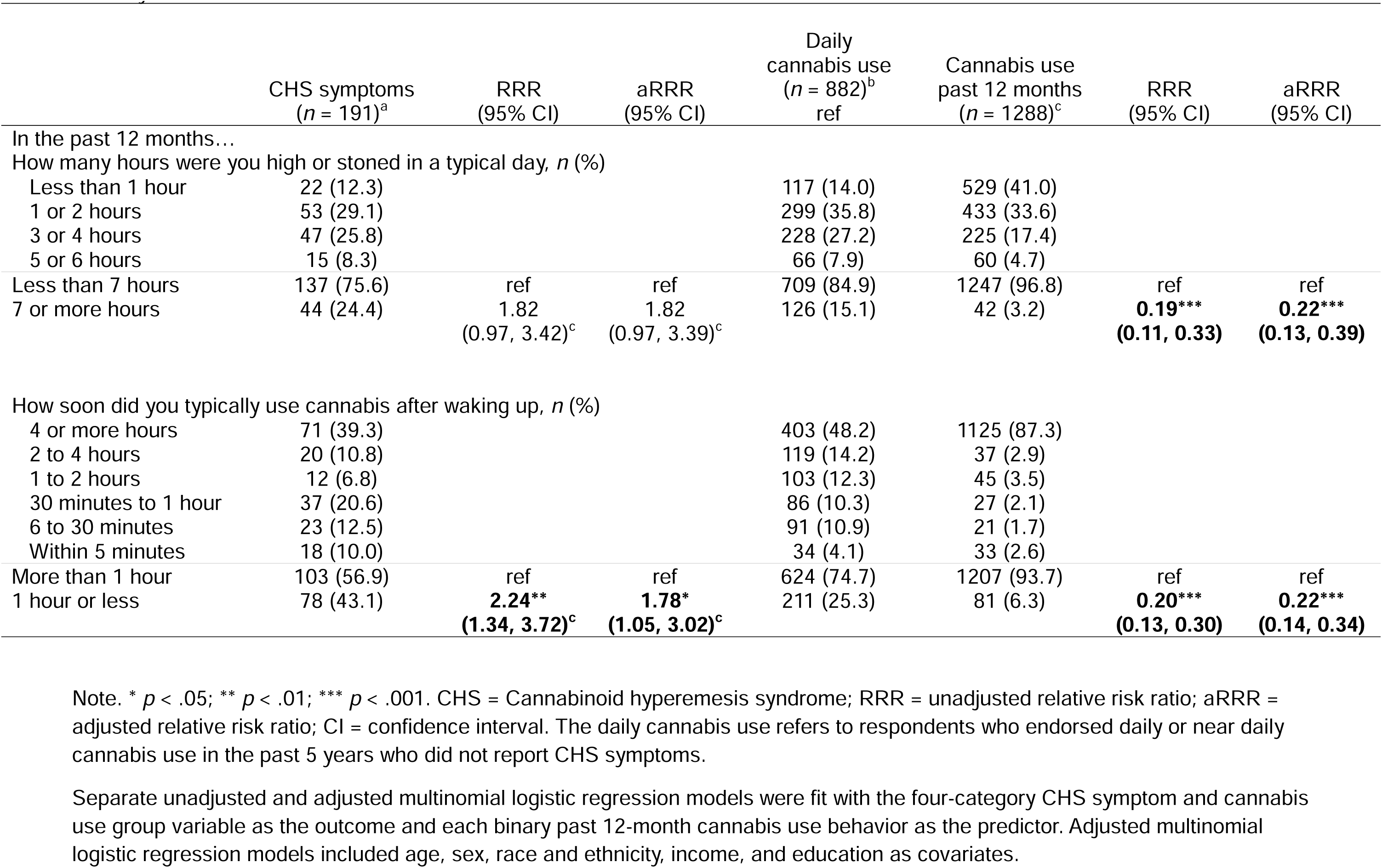
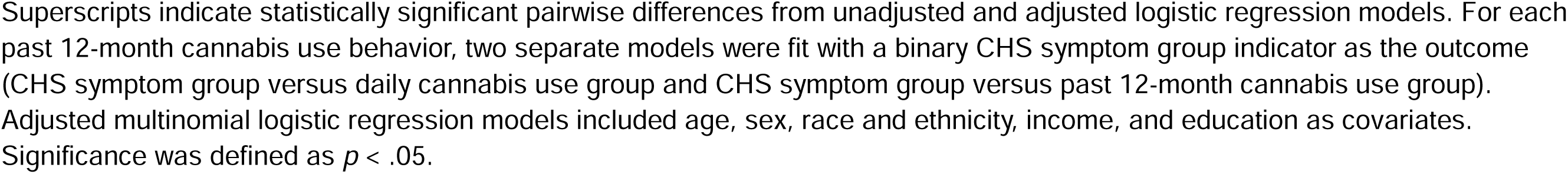
Past 12-month cannabis use behaviors among CHS symptom and cannabis use groups; National Firearms, Alcohol, Cannabis, and suicide survey, 2025.

The prevalence of using cannabis within 1 hour of waking was 43.1% in the CHS symptom group, compared with 25.3% in the daily cannabis use group and 6.3% in the cannabis use in the past 12-months group. In both unadjusted and adjusted models, using cannabis within 1 hour of waking was significantly associated with a higher likelihood of being in the CHS symptom group and lower likelihood of being in the cannabis use in the past 12-month group, relative to the daily use group. In post hoc logistic regression models, the CHS symptom group was more likely to report using cannabis within 1 hour of waking than the past 12-month cannabis use group.

### Cannabis problem use

The lifetime prevalence of each cannabis use problem was highest among those in the CHS symptom group compared to those in the other cannabis use groups (see Table 4). In unadjusted models, all lifetime cannabis use problems except use in situations that could be physically hazardous were significantly associated with a greater likelihood of being in the CHS symptom group relative to the daily cannabis use group. After covariate adjustment, endorsement of each cannabis use-related problem was significantly associated with greater likelihood of being in the CHS symptom group, and lower likelihood of being in the cannabis use in the past 12-months or no cannabis use in the past 12-months groups, relative to the daily use group. Similarly, higher continuous lifetime cannabis problem use scores were significantly associated with greater likelihood of being in the CHS symptom group and lower likelihood of being in the cannabis use in the past 12-months and no cannabis use in the past 12-months groups compared to the daily cannabis use group. Among the cannabis-related problems examined, continued use despite knowing it caused or exacerbated problems, devoting a great of time to cannabis use, and failing to meet expectations due to cannabis were most strongly associated with being the CHS symptom group versus the daily use group. In unadjusted and adjusted post hoc logistic regression models, endorsement of each lifetime cannabis use problem significantly differed between the CHS symptom group relative to the past 12-month cannabis use and no cannabis use groups. A similar pattern of results was observed for past 12-month cannabis use problems (see eTable 1 in the Supplement).

**Table 4.**
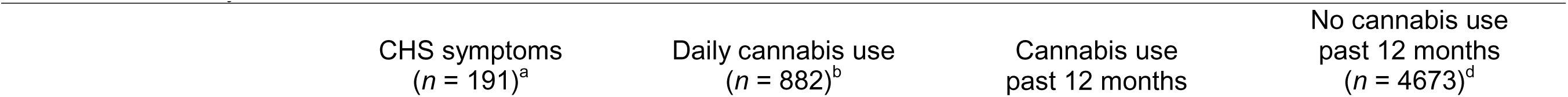
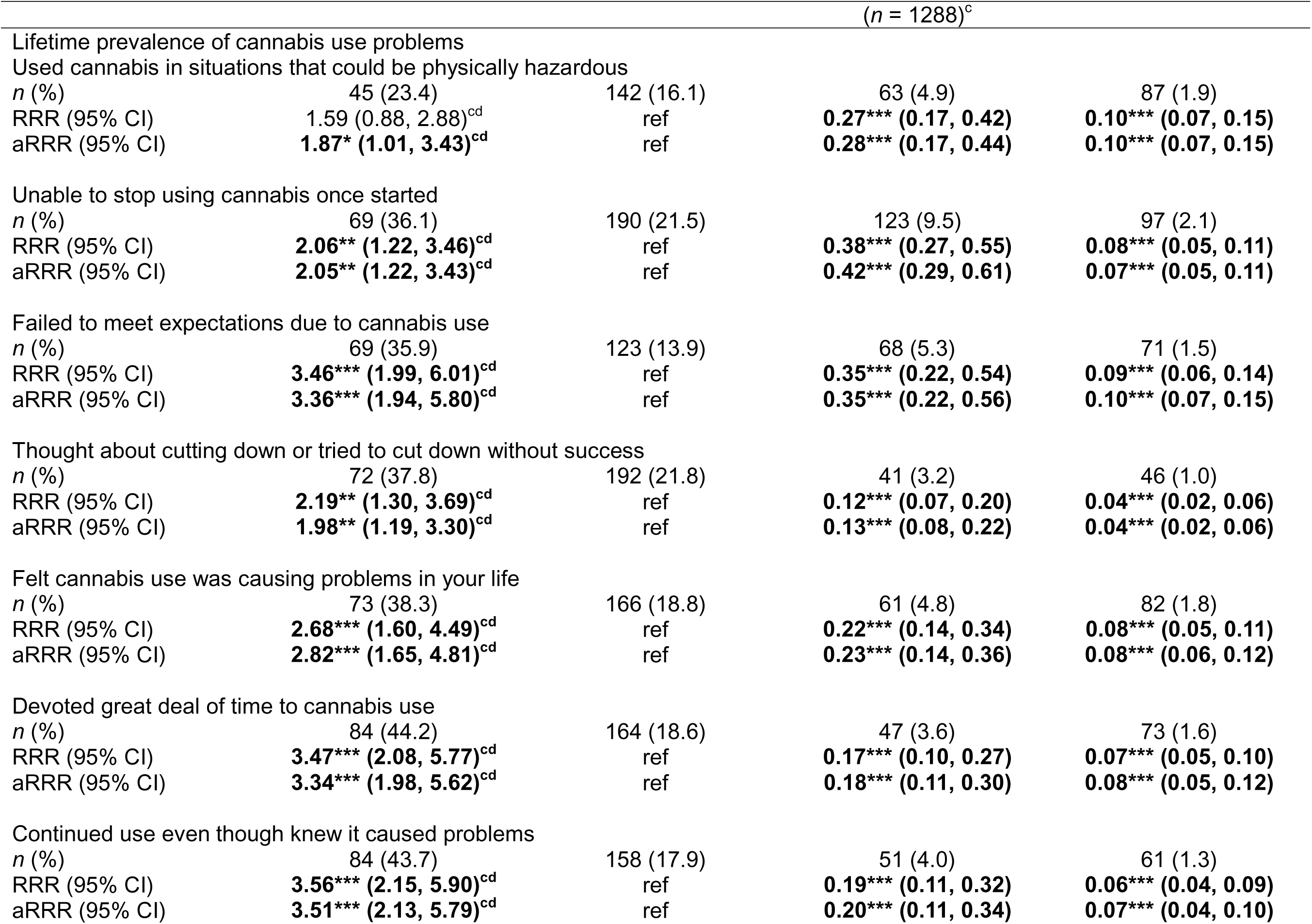
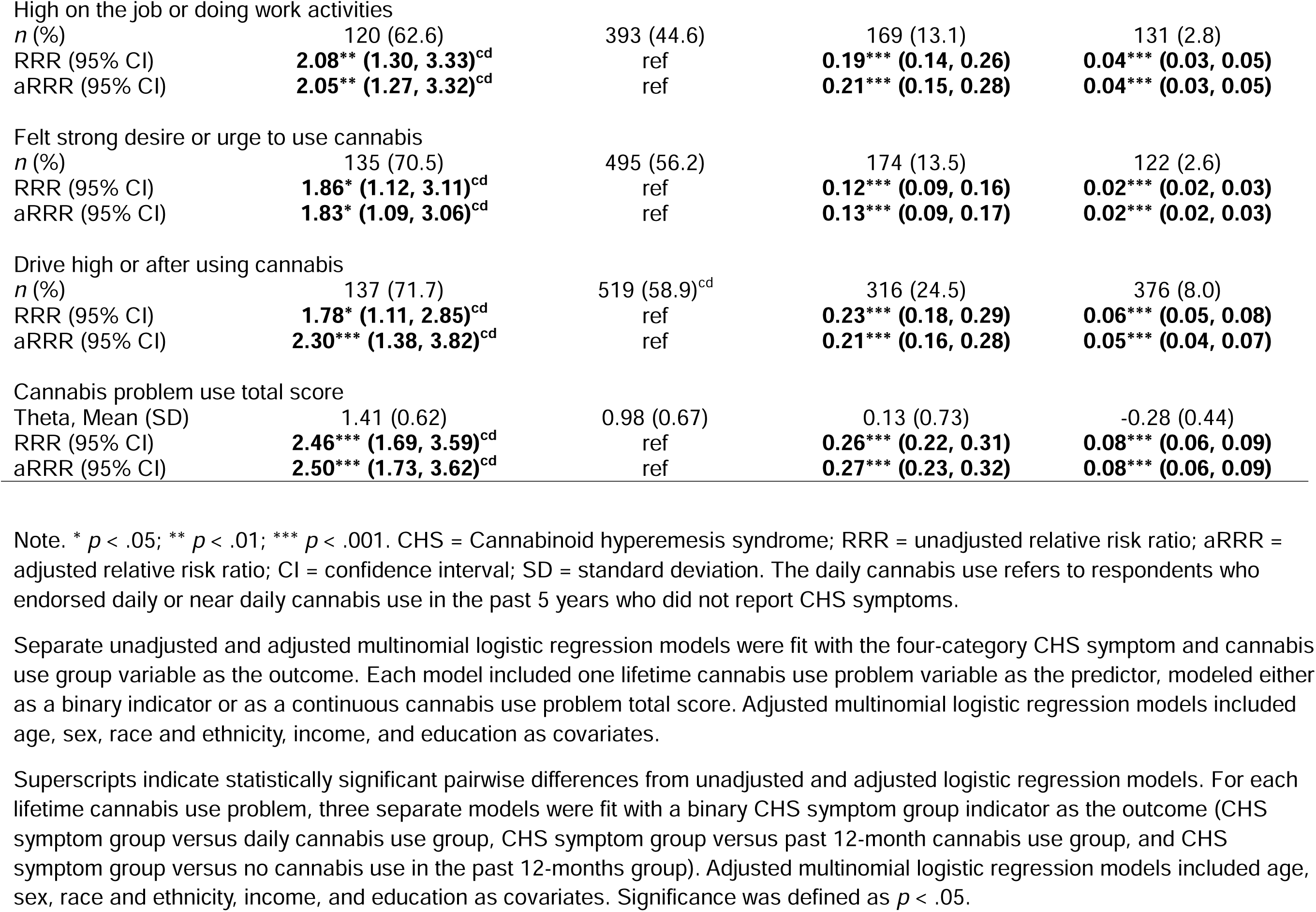
Lifetime cannabis problem use among CHS symptom and cannabis use groups; National Firearms, Alcohol, Cannabis, and Suicide survey, 2025.

## Discussion

As frequent cannabis use continues to rise,^1, 2^ and the US pursues reclassifying cannabis under the Controlled Substances Act (CSA) from a Schedule I to Schedule III substance,^28^ it is increasingly important ascertain the potential public health implications of regular cannabis use among a growing portion of US citizens. CHS has recently been described in the clinical literature as a condition characterized by repeated episodes of severe nausea and vomiting among individuals who use cannabis regularly.^5–10^ Our findings indicate that a significant proportion of those who use cannabis daily report symptoms consistent with CHS, with nearly 18% of those who used cannabis daily in the past five years endorsing these symptoms, translating to over seven million US adults reporting symptoms that could be related to CHS and a little over a million US adults reporting that they have been diagnosed by a medical provider with CHS. The majority of those with potential symptoms of CHS reported that they have not been diagnosed with CHS by a medical provider, suggesting that previous clinical estimates may have underestimated the prevalence of CHS and that significant gaps in care exist for those potentially struggling with CHS.

This survey included items intended to map onto the description of CHS that exists in clinical literature,^5–10^ but have not been validated or compared to consensus clinical diagnosis. However, several findings in this survey add credibility to the potential value of these initial items to capture potential symptoms of CHS. Consistent with the clinical literature, 47.2% reported that their symptoms improved following a hot bath or shower and 27.6% reported symptom improvement after stopping cannabis.^9, 13, 18, 19^ Unfortunately, the wording of the questions did not allow for the determination of whether these strategies were attempted and did not work, versus were not tried. Subsequent comparisons indicated that CHS symptoms were associated with more frequent cannabis use, use earlier in the day, more hours of the day spent “high”, and more cannabis use problems (similar to symptoms of cannabis use disorder) compared to those without any cannabis use or less than daily cannabis use. Most importantly, the CHS symptom group exhibited more cannabis use problems than others who used cannabis daily, consistent with the notion the CHS symptom group is a unique group even among those who use cannabis frequently. These results are consistent with a prior survey of attendees of online support groups for CHS which found that 82.2% reported cannabis use 3 or more times daily before the onset of CHS.^29^ Although some of the differences were small, the CHS symptom group was also younger, more likely to female and non-White race, lower income, and less educated than the other cannabis use groups.

A challenge to characterizing individuals with CHS is the lack of clear, consensus diagnostic criteria. Many of the commonly used diagnostic frameworks such as the Rome IV criteria^30^ rely upon relief of nausea and vomiting episodes after sustained abstinence in order to make the diagnosis. The requirement of sustained cessation of cannabis use prior to conferring CHS diagnosis presents multiple challenges, however, as individuals may not perceive their symptoms to be related to their cannabis use,^31^ they may not have attempted to abstain from cannabis use. Additionally, many diagnostic frameworks require a certain duration of symptoms such as 6 months prior to diagnosis, which can contribute to under-recognition and underdiagnosis of initial episodes. The current survey items aimed to be more inclusive of people with CHS symptoms who are at various stages in the syndrome and who may not have quit or attempted to quit cannabis use.

There is also a lack of consensus regarding treatment guidelines for CHS, although there are emerging clinical guidelines and frameworks.^15, 32, 33^ However, the existing literature is focused on the care administered in acute care settings and strategies to decrease re-admission, and not on the experiences of the majority of people with CHS who are in outpatient settings such as primary care clinics and addiction treatment programs, or those who have not sought medical care for their symptoms and may be better reached via public health or community-based initiatives.

This study had several limitations related to the measurement of CHS symptoms. First, because the CHS symptom items were a small part of a much larger survey, the items were limited in terms of details regarding the history, duration, and intensity of symptoms so the items cannot distinguish clinically significant CHS from milder CHS symptoms that are associated with minimal functional impairment. Second, alternative conditions could account for some of the episodes of nausea and vomiting. This last limitation is important because the definition of CHS in the present study may include individuals who were misclassified as having CHS symptoms but for whom there was no causal link between the cannabis use and gastrointestinal symptoms, or even reverse causation whereby cannabis was used in an attempt to relieve gastrointestinal symptoms (e.g., cannabis used for symptom relief from cancer chemotherapy). Third, the CHS items were generated de novo for this survey and have their own limitations. For example, the goal of asking about a period of daily or near daily cannabis use in the past 5 years was to obtain a group with a period of frequent cannabis use that could lead CHS. However, this may not be the optimal way of operationalizing heavy use that could lead to CHS. The criteria related to improvement in symptoms after hot baths/showers or cessation of cannabis use do not ask about whether these strategies were attempted and led to symptom improvement or were not attempted at all. These limitations with the measurement of CHS symptoms clearly decrease the precision of the measurement of CHS symptoms; however, because there are no prior attempts to capture the national prevalence of CHS symptoms, this work provides a starting point from which the field can build more precise estimates.

Although the overall sample was large, smaller cell sizes in certain groups limit the precision of estimates and the extent to which multivariable analyses can be conducted. There are general limitations associated with the survey design including the need for online access and proficiency in English to complete the survey, and respondents may have misinterpreted certain questions. Selection bias could have also occurred whereby the people who responded to the survey differed from those who did not in systematic but unmeasured ways. For the present study, it is possible that those with severe CHS were not willing or able to complete the survey due to the severity of their symptoms; thus, underestimating the extent of CHS symptoms in the population. While the survey weights help to better approximate the target population, their statistical adjustments are constrained to a set of measured variables and so do not adjust the sample observations for any bias in other measured or unmeasured variables or for any potential non-response bias.

## Public Health Implications

In 2022, for the first time, the number of individuals who use cannabis daily or near-daily surpassed the number of those who used alcohol daily or near-daily^3^ and the consequences of these shifts in behavior are just beginning to be understood. Results from this national survey indicate that many US adults endorse symptoms of CHS, indicating that this could be an emerging consequence of daily cannabis use. Future work is needed to better understand the risk factors for developing CHS, novel settings for care, and potential treatments for CHS.

## Data Availability

All data in this article will become publicly available in the National Data Archive managed by the National Institute of Health at the end of the grant period.

https://osf.io/7w4ns/overview

