## supplemental table 1 for "Prevalence and Correlates of Symptoms of Cannabinoid Hyperemesis Syndrome in the United States"

**Supplemental Results – eTable 1**

*Past 12-month cannabis problem use*. eTable 1 presents prevalence estimates and multinomial logistic regression results for past 12-month cannabis use problems across the CHS symptom and cannabis use groups. As in the primary analyses, we fit separate unadjusted and adjusted multinomial logistic regression models, with the four-category CHS symptom and cannabis group variable as the outcome and each past 12-month cannabis use problem as the predictor, modeled either as a binary indicator or as the continuous cannabis use problem total score. Adjusted models included age, sex, race and ethnicity, income, and education as covariates. In post hoc analyses, we fit separate unadjusted and adjusted logistic regression models using binary group indicators as the outcomes to assess pairwise differences between the CHS symptom group and other cannabis use groups on past 12-month cannabis use problems.

Endorsement of each cannabis use problem was more prevalent in the CHS symptom and daily cannabis groups than in the cannabis use in the past 12-months group. In unadjusted models, all past 12-month cannabis use problems except use in situations that could be physically hazardous and driving while high or after cannabis use were significantly associated with greater likelihood of being in the CHS symptom group versus the daily cannabis use group. After covariate adjustment, the associations of being high on the job or when doing work activities and feeling a strong desire or urge to use cannabis with being in the CHS symptom group, relative to the daily cannabis use group, were slightly attenuated and no longer significant. Among the past 12-month cannabis-related problems examined, continued use despite knowing it caused or exacerbated problems, devoting a great of time to cannabis use, and failing to meet expectations due to cannabis were most strongly associated with being the CHS symptom group versus the daily use group in unadjusted and adjusted models. Across unadjusted and adjusted models, each cannabis use-related problem was significantly associated with lower likelihood of being in the cannabis use in the past 12-months group, relative to the daily use group. Higher continuous past 12-month cannabis problem use scores were also significantly associated with greater likelihood of being in the CHS symptom group and lower likelihood of being in the cannabis use in the past 12-month group relative to the daily use group. Unadjusted and adjusted post hoc logistic regression indicated that endorsement of each past 12-month cannabis use problem significantly differed between the CHS symptom group and the past 12-month cannabis use group.

| eTable 1. Past 12-month cannabis problem use among CHS symptom and cannabis use groups | | | |
| --- | --- | --- | --- |
|  | CHS symptoms  (*n* = 191)^a^ | Daily cannabis use  (*n* = 882)^b^ | Cannabis use  past 12 months  (*n* = 1288)^c^ |
| Past 12-month prevalence of cannabis use problems | | | |
| Used cannabis in situations that could be physically hazardous | | | |
| *n* (%) | 30 (15.5) | 88 (10.0) | 34 (2.6) |
| RRR (95% CI) | 1.66 (0.81, 3.43)^c^ | ref | **0.24*** (0.13, 0.47)** |
| aRRR (95% CI) | 1.69 (0.81, 3.53)^c^ | ref | **0.26*** (0.13, 0.52)** |
| Unable to stop using cannabis once started | | | |
| *n* (%) | 46 (24.2) | 79 (8.9) | 32 (2.5) |
| RRR (95% CI) | **3.25*** (1.67, 6.34)^c^** | ref | **0.26*** (0.14, 0.49)** |
| aRRR (95% CI) | **2.59** (1.31, 5.10)^c^** | ref | **0.30*** (0.15, 0.58)** |
| Failed to meet expectations due to cannabis use | | | |
| *n* (%) | 49 (25.5) | 69 (7.8) | 44 (3.4) |
| RRR (95% CI) | **4.05*** (2.12, 7.74)^c^** | ref | **0.42** (0.24, 0.74)** |
| aRRR (95% CI) | **3.54*** (1.85, 6.77)^c^** | ref | **0.43** (0.23, 0.78)** |
| Thought about cutting down or tried to cut down without success | | | |
| *n* (%) | 61 (31.8) | 144 (16.4) | 23 (1.8) |
| RRR (95% CI) | **2.39** (1.35, 4.22)^c^** | ref | **0.09*** (0.05, 0.18)** |
| aRRR (95% CI) | **1.97* (1.12, 3.47)^c^** | ref | **0.10*** (0.05, 0.19)** |
| Felt cannabis use was causing problems in your life | | | |
| *n* (%) | 61 (32.0) | 120 (13.6) | 31 (2.4) |
| RRR (95% CI) | **2.98*** (1.71, 5.20)^c^** | ref | **0.16*** (0.09, 0.29)** |
| aRRR (95% CI) | **2.80*** (1.56, 5.04)^c^** | ref | **0.16*** (0.09, 0.30)** |
| Devoted great deal of time to cannabis use | | | |
| *n* (%) | 66 (34.8) | 98 (11.2) | 22 (1.7) |
| RRR (95% CI) | **4.24*** (2.38, 7.56)^c^** | ref | **0.14*** (0.07, 0.28)** |
| aRRR (95% CI) | **3.55*** (1.97, 6.42)^c^** | ref | **0.15*** (0.07, 0.30)** |
| Continued use even though knew it caused problems | | | |
| *n* (%) | 69 (36.3) | 113 (12.8) | 36 (2.8) |
| RRR (95% CI) | **3.87*** (2.24, 6.69)^c^** | ref | **0.20*** (0.10, 0.37)** |
| aRRR (95% CI) | **3.53*** (2.02, 6.18)^c^** | ref | **0.21*** (0.11, 0.40)** |
| High on the job or doing work activities | | | |
| *n* (%) | 77 (40.1) | 253 (28.7) | 43 (3.3) |
| RRR (95% CI) | **1.66* (1.02, 2.73)^c^** | ref | **0.08*** (0.05, 0.14)** |
| aRRR (95% CI) | 1.44 (0.87, 2.38)^c^ | ref | **0.09*** (0.06, 0.15)** |
| Felt strong desire or urge to use cannabis | | | |
| *n* (%) | 121 (63.3) | 450 (51.0) | 142 (11.0) |
| RRR (95% CI) | **1.66* (1.03, 2.67)^c^** | ref | **0.12*** (0.09, 0.16)** |
| aRRR (95% CI) | 1.48 (0.91, 2.41)^c^ | ref | **0.12*** (0.09, 0.17)** |
| Drive high or after using cannabis | | | |
| *n* (%) | 97 (50.6) | 373 (42.3) | 146 (11.3) |
| RRR (95% CI) | 1.40 (0.88, 2.22)^c^ | ref | **0.17*** (0.13, 0.24)** |
| aRRR (95% CI) | 1.44 (0.91, 2.27)^c^ | ref | **0.18*** (0.13, 0.24)** |
| Cannabis problem use total score | | | |
| Theta, Mean (SD) | 0.98 (0.88) | 0.50 (0.73) | -0.32 (0.56) |
| RRR (95% CI) | **2.03*** (1.50, 2.75)^c^** | ref | **0.17*** (0.14, 0.23)** |
| aRRR (95% CI) | **1.91*** (1.40, 2.61)^c^** | ref | **0.17*** (0.14, 0.23)** |

Note. * *p* < .05; ** *p* < .01; *** *p* < .001. CHS = Cannabinoid hyperemesis syndrome; RRR = unadjusted relative risk ratio; aRRR = adjusted relative risk ratio; CI = confidence interval; SD = Standard deviation. The daily cannabis use refers to respondents who endorsed daily or near daily cannabis use in the past 5 years who did not report CHS symptoms.

Separate unadjusted and adjusted multinomial logistic regression models were fit with the four-category CHS symptom and cannabis use group variable as the outcome. Each model included one past 12-month cannabis use problem variable as the predictor, modeled either as a binary indicator or as the continuous cannabis use problem total score. Adjusted multinomial logistic regression models included age, sex, race and ethnicity, income, and education as covariates.

Superscripts indicate statistically significant pairwise differences from unadjusted and adjusted logistic regression models. For each past 12-month cannabis use problem, two separate models were fit with a binary CHS symptom group indicator as the outcome (CHS symptom group versus daily cannabis use group and CHS symptom group versus past 12-month cannabis use group). Adjusted logistic regression models included age, sex, race and ethnicity, income, and education as covariates. Significance was defined as *p* < .05.
